# Homologous and Variant-Specific Memory B-Cell and Antibody Responses after SARS-CoV-2 mRNA Vaccination

**DOI:** 10.1101/2021.07.12.21260386

**Authors:** Iana H. Haralambieva, Jonathon M. Monroe, Inna G. Ovsyannikova, Diane E. Grill, Gregory A. Poland, Richard B. Kennedy

## Abstract

**Importance:** A better understanding of the immune memory and functional humoral immunity directed at the emerging Variants of Concern (VoC) strains after SARS-CoV-2 vaccination is essential for predicting the longevity of heterotypic protection.

**Objective:** The aim of our study was to characterize functional humoral immunity (including memory B cell response) after COVID-19 mRNA vaccination and to determine/compare the reactivity of COVID-19 vaccine-induced memory B cells to the emerging SARS-CoV-2 Variants of Concern (VoC).

**Design, setting, participants and interventions:** We designed an exploratory longitudinal observational (convenience sample-based) study at Mayo Clinic, Rochester, MN that enrolled and followed naïve subjects and recovered COVID-19 subjects from Olmsted County, MN and surrounding areas after COVID-19 vaccination in January-June 2021. The study enrolled 17 relatively healthy subjects, 59% females and 94% White/Non-Hispanic or Latino with median age at enrollment 41 years. The subjects received either the BNT162b2 (Pfizer/BioNtech) or mRNA-1273 (Moderna) vaccine (n=3) and provided a blood sample at baseline, at ∼3 weeks after their first vaccine dose/before the second dose, and at ∼2 weeks after the receipt of their second vaccine dose.

**Main outcomes and measures:** Spike-specific humoral and memory B cells responses were assessed over time after vaccination against the original Wuhan-Hu-1/vaccine and against emerging VoC strains/antigens.

**Results:** We observed a robust neutralizing antibody response after COVID-19 mRNA vaccination, but a reduction in the functional antibody activity to several of the emerging SARS-CoV-2 VoC. Consistent with this, we also found differences in the number of isotype-switched/IgG+ MBCs responding to homologous and variant receptor-binding domain/RBDs after vaccination. We found a reduction of MBCs reactive to RBDs of Beta, Gamma and Delta SARS-CoV-2 VoC strains.

**Conclusion and relevance:** In this exploratory study in subjects following receipt of COVID-19 mRNA vaccine, we found differences in antibody titers observed for VoCs after vaccination that are accompanied with, and can partially be explained by, decreased MBC reactivity against the VoCs. This can further attenuate the generated recall humoral immune response upon exposure to these variants.

**Key Points:** *Question:* What is the reactivity of COVID-19 vaccine-induced memory B cells to the emerging SARS-CoV-2 Variants of Concern (VoC)?

*Findings:* In this longitudinal cohort study of subjects receiving COVID-19 mRNA vaccine we assessed memory B cell response and functional antibody titers. We found statistically significant differences between the frequencies of memory B cells responding to homologous and VoC receptor-binding domain/RBDs after vaccination.

*Meaning:* In concert with the lowered antibody response, the reduced memory B-cell response to VoC could translate to an increased susceptibility to the emerging SARS-CoV-2 variant strains in the face of waning immunity.

## Introduction

The longevity of protective humoral immunity after SARS-CoV-2 infection and vaccination is largely dependent on the generation and persistence of antigen-specific isotype-switched memory B cells (MBCs), as well as long-lived plasma cells residing in the bone marrow. IgG is considered the dominant isotype of MBCs after COVID-19 infection^1^, and the frequency of receptor binding domain/RBD-specific IgG+ MBCs is considered to represent a marker of immune memory following SARS-CoV-2 infection/vaccination.^2,3^ The reactivity of mRNA vaccine-induced MBCs to emerging clinically significant SARS-CoV-2 variant of concern (VoC) strains is unknown and was assessed in this study in concert with functional antibody measures.

## Methods

### Human Subjects

The study sample consisted of 17 healthy subjects (15 naïve and 2 recovered from COVID-19 infection) who provided a blood sample prior to vaccination (baseline) with BNT162b2 (Pfizer/BioNtech) or mRNA-1273 (Moderna) vaccine, then at ∼3 weeks after their first vaccine dose and before the second dose (first vaccine dose timepoint), and at ∼2 weeks after the receipt of their second vaccine dose (second vaccine dose timepoint). All study participants provided written informed consent, and all study procedures were approved by the Mayo Clinic Institutional Review Board.

### Antibody Assays

The neutralizing antibody response was assessed using a high-throughput, fluorescence/GFP-based pseudovirus/VSV microneutralization assay^4^, further developed in our laboratory to allow rapid imaging/quantification of cell infection with the ImageXpress® platform/software (Molecular Devices). Wuhan-Hu-1 Spike-specific 50% end-point titer (Neutralizing Dose, ND_50_) for each sample was calculated using Karber’s formula. The capacity of sera (post-second vaccination) to impair virus attachment/block the interaction between hACE2 and different RBDs was determined using the SARS-CoV-2 Surrogate Virus Neutralization Test/sVNT Kit (RUO.3.0, GenScript® USA). The dilution at which a sample crosses 50% inhibition is designated as the 50% inhibition dose (ID_50_).

### MBC ELISPOT Assay

The frequencies of SARS-CoV-2-specific MBCs were quantified using the Mabtech ELIspot^PLUS^ kit for human IgG (Mabtech Inc.; Cincinnati, OH) using recombinant SARS-CoV-2 antigens (Sino Biologicals, Beijing, China) coated at 0.2 µg/well. Responses are presented in spot-forming units (SFUs) per 2×10^5^ cells.

### Statistical Analysis

The Wilcoxon signed-rank test was used to assess differences for all comparisons. The ID_50_ value was calculated for each subject and RBD by fitting a quadratic regression of the percent inhibition on the log_3_ inverse dilution. The quadratic equation utilizing the regression coefficients was used to calculate the inverse dilution from the regression equation at 50% inhibition.

## Results

Our study cohort consisted of 59% females and 94% White/Non-Hispanic or Latino. Their median age at enrollment was 41 years (IQR 24, 54). The majority of subjects received the Pfizer BNT162b2 and three subjects received the Moderna mRNA-1273 vaccine series, both according to the standard schedule.

### Antibody Response

As expected, SARS-CoV-2 vaccination elicited robust anti-Spike neutralizing antibody response with significant inter-individual variations. Titers increased significantly after each vaccine dose (p-value < 0.001 for all comparisons, Fig. 1A). The median ND_50_ at baseline was 27 (excluding one subject with documented COVID-19 before the enrollment with a baseline ND_50_ of 225). The median ND_50_ at the peak of the humoral immune response, at ∼ 2 weeks after the second SARS-CoV-2 vaccination, was 3,290 (IQR 2,572; 4,018), which is consistent with other studies reporting a significant boost in functional humoral immunity upon completion of the vaccination series (Fig. 1A). Analysis of the functional ability of sera from subjects to inhibit/block the RBD-ACE2 binding for three VoC demonstrated a reduction in function (more pronounced for the Beta/RBD B.1.351 and the Gamma/RBD P.1 variants) compared to the inhibition of the Wuhan-Hu-1 RBD-hACE2 binding (Fig. 1B). The calculated median ID_50_ values for the tested RBD antigens were 1,200 for the Wuhan-Hu-1 RBD), and 498, 325 and 224 for the Alpha, Beta and Gamma RBDs, respectively.

**Fig. 1.**
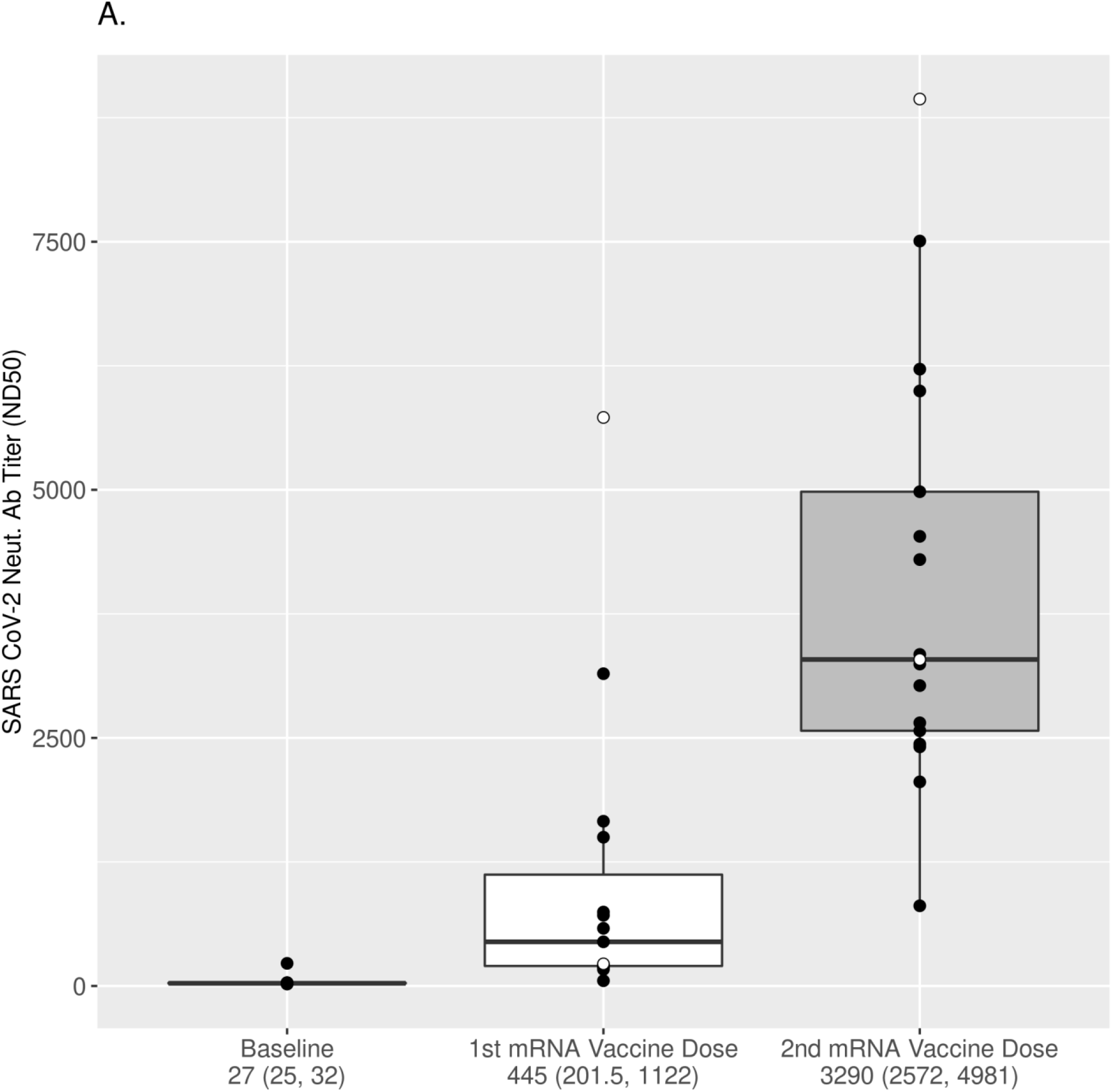
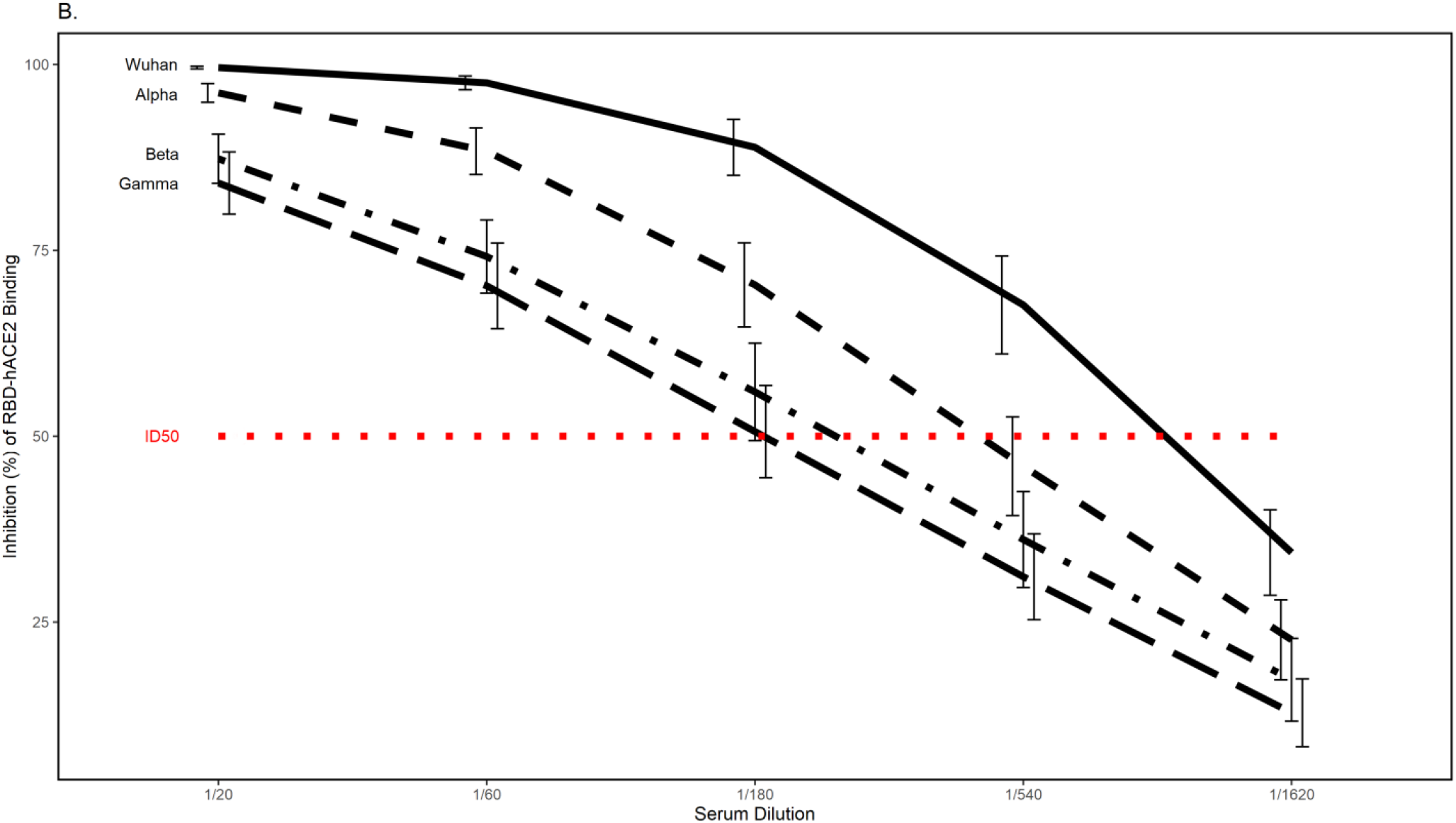
Functional antibody response after COVID-19 mRNA vaccination. (A) The dynamics of neutralizing antibody response after vaccination was assessed using a pseudovirus microneutralization assay (see Methods). ND_50_ values over three time points relative to vaccination are displayed using boxplots for all subjects. The values are plotted in log2 scale, but the scales of the axis reflect the untransformed values for easier interpretation. Each box was plotted using the 25% to 75% interquartile range and the median is represented by the bold line in the box. The “ whiskers” extend up to 1.5 times the interquartile range above or below the 75th or 25th percentiles, respectively. Black dots represent naïve (at baseline) subjects, while white dots represent COVID-19-recovered (at baseline) subjects. The numbers below the x-axis represent the median response for all subjects (25% and 75% interquartile range). Titers increased significantly after each vaccine dose (p-value<0.001 for all comparisons, Wilcoxon signed-rank test). (B) Comparison of serum Ab-mediated inhibition of hACE2–RBD binding (n=8, after second vaccine dose) for different RBDs of VoC (Alpha, Beta, Gamma) and the Wuhan-Hu-1 RBD using the GenScript sVNT kit. Samples were serially diluted (3-fold) from 1/20 to 1/1620 and the different dilutions were assessed for blockade/inhibition of RBD-hACE2 interaction. The dashed red line represents 50% reduction in RBD-hACE2 binding compared to the negative controls (absence of blocking antibodies). ID_50_ values to different RBDs were calculated as described in Methods. ID_50_ values to Wuhan-Hu-1 RBD are significantly greater (p=0.008, Wilcoxon signed rank-test) than the ID_50_ values of each VoC RBD. Error bars represent standard error.

### MBC Response

We profiled the frequencies and specificities of isotype-switched IgG+ MBC after SARS-CoV-2 mRNA vaccination, as an indication of mature, highly specific and functional immune memory. As expected, the antigen-specific MBC changed significantly over time during mRNA vaccination (Fig. 2A, p-value<0.006 for all comparisons between timepoints for S1 and RBD MBC). The prime vaccination elicited significant SARS-CoV-2-specific MBC response directed mainly to the S1 portion of the SARS-CoV-2 Spike of Wuhan-Hu-1and particularly to its RBD (Fig. 2A). This response was further boosted after the second vaccine dose to a median S1 MBC response of 45 SFUs per 2×10^5^ cells and a median RBD response of 27 SFUs per 2×10^5^ cells (Fig. 2A). Interestingly, IgG+ MBC directed to the S2 potion of the Spike, as well as to the N-terminal domain/NTD portion of the S1 were readily detectable for some of the subjects, but at a much lower frequency (Fig. 2B). We also noted moderate, but consistent correlations between the ND_50_ titers and the frequency of S1-and RBD-specific MBCs after first vaccine dose (r=0.82 for both, p-value=0.00019 and p=0.0002, respectively). The correlations were less robust with marginal statistical significance after the second vaccine dose (r=0.49/r=0.43, p-value=0.046 and p=0.08, respectively); likely due to the small sample size and inter-individual variability in vaccine response.

**Fig. 2.**
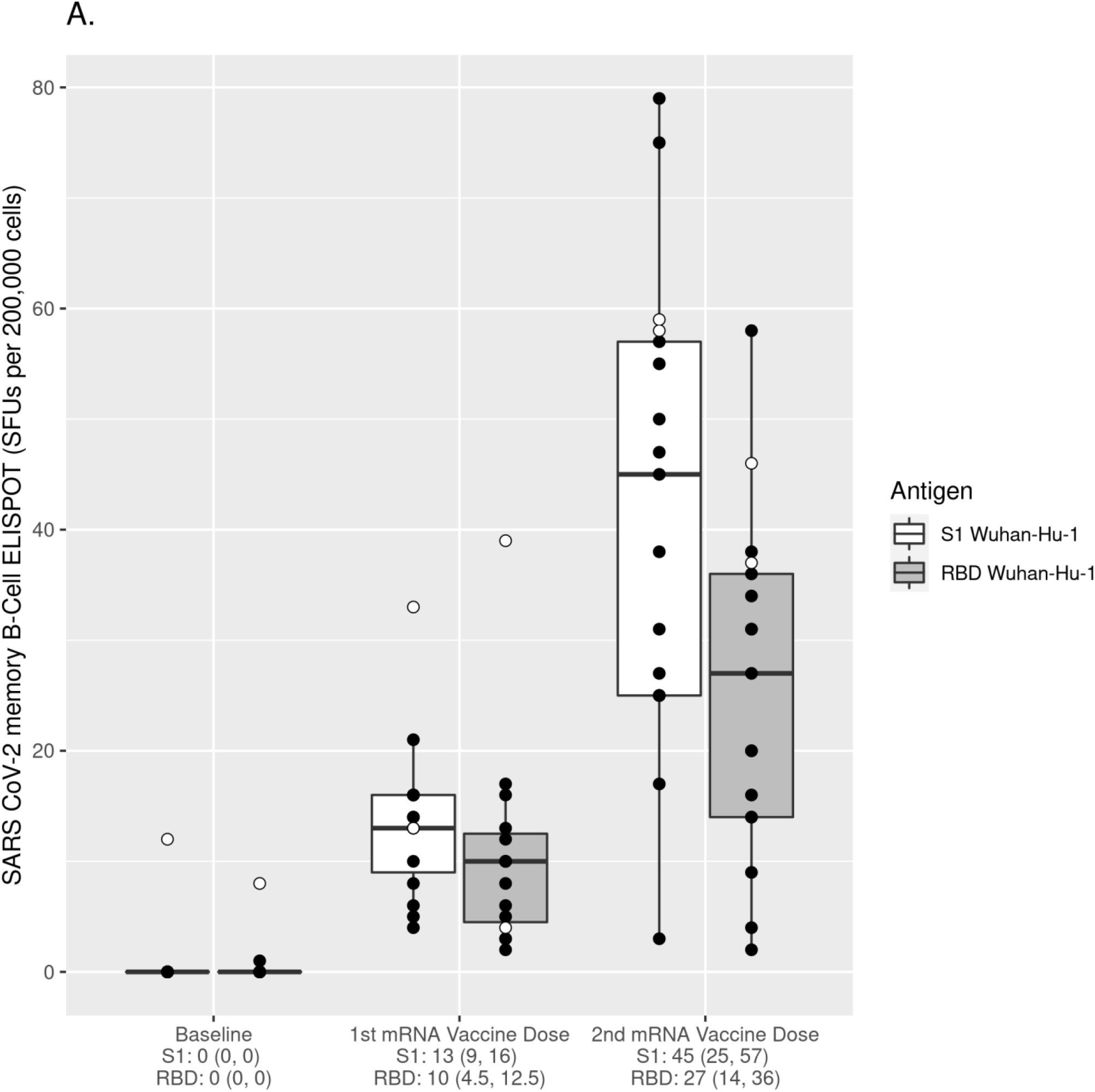
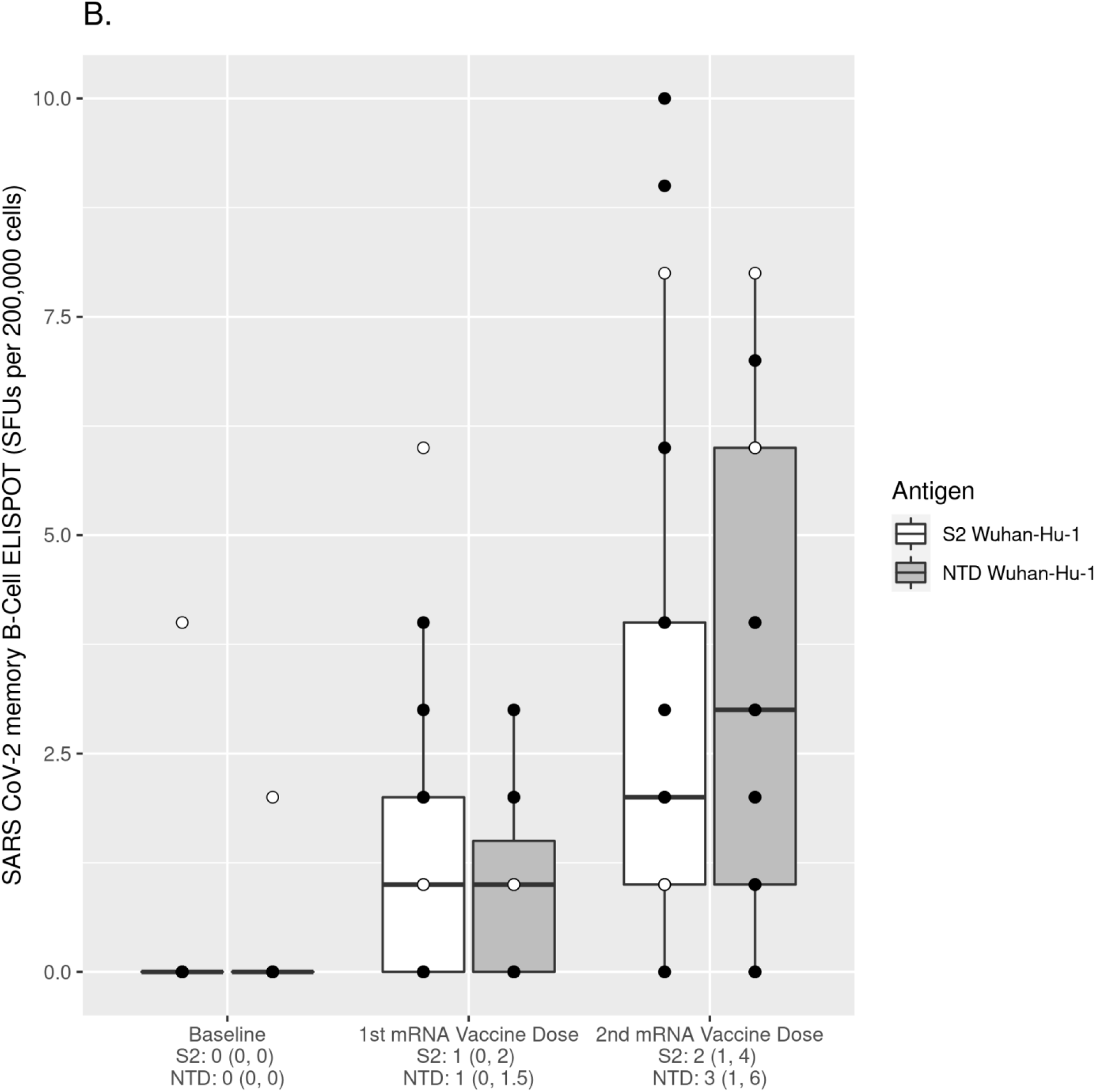
Dynamics of MBC response after COVID-19 mRNA vaccination. The frequencies of IgG+ MBCs were measured using the Mabtech ELIspot^PLUS^ kit for human IgG using the indicated antigens (dark grey and white boxes), after *in vitro* PBMC three-day stimulation with human recombinant IL-2 and R848. Detected responses are presented in spot-forming units (SFUs) per 2×10^5^ cells, as subjects’ antigen-specific medians (from three replicates with subtracted subject-specific no-antigen background measure). The values are plotted in log2 scale, but the scales of the axis reflect the untransformed values for easier interpretation. Each box was plotted using the 25% to 75% interquartile range and the median was represented by the bold line in the box. The “ whiskers” extend up to 1.5 times the interquartile range above or below the 75th or 25th percentiles, respectively. Black circles represent naïve subjects (at baseline), while white circles represent COVID-19-recovered subjects (at baseline). The numbers below the x-axis represent the median response for all subjects (25% and 75% interquartile range). The MBC responses and changes over time of S1/RBD-specific MBC response (A) and S2/NTD MBC response (B) are assessed using the Wilcoxon signed-rank test. (A) S1/RBD-specific MBC response: p<0.006 for all comparisons (first dose vs. baseline, second dose vs baseline, and second dose vs first dose) and (B) S2/NTD MBC response: p<0.05 for all comparisons. Please, note the different scale on the two graphs.

### Differential Reactivity of MBCs to VoC RBDs

We then probed the reactivity/ability of the IgG+ RBD-specific MBCs to target RBDs of emerging, clinically significant VoC strains (Alpha/RBD B.1.1.7, Beta/RBD B.1.351, Gamma/RBD P.1 and Delta/RBD B.1.617) and the results are displayed in Fig. 3. While RBD-specific MBCs were readily detectable to all variant RBDs at both timepoints following mRNA vaccination, we noted statistically significant reductions in the MBC frequencies to the each of the variant RBDs after the first vaccine dose (p<0.025 for all comparisons, Fig. 3A). Following the second vaccine dose, a significant decrease in the number of reactive MBCs recognizing the Beta, Gamma and Delta RBD variants remained (p<0.002 for all comparisons), while the difference in MBC response to the Alpha variant was non-significant (Fig. 3B).

**Fig. 3.**
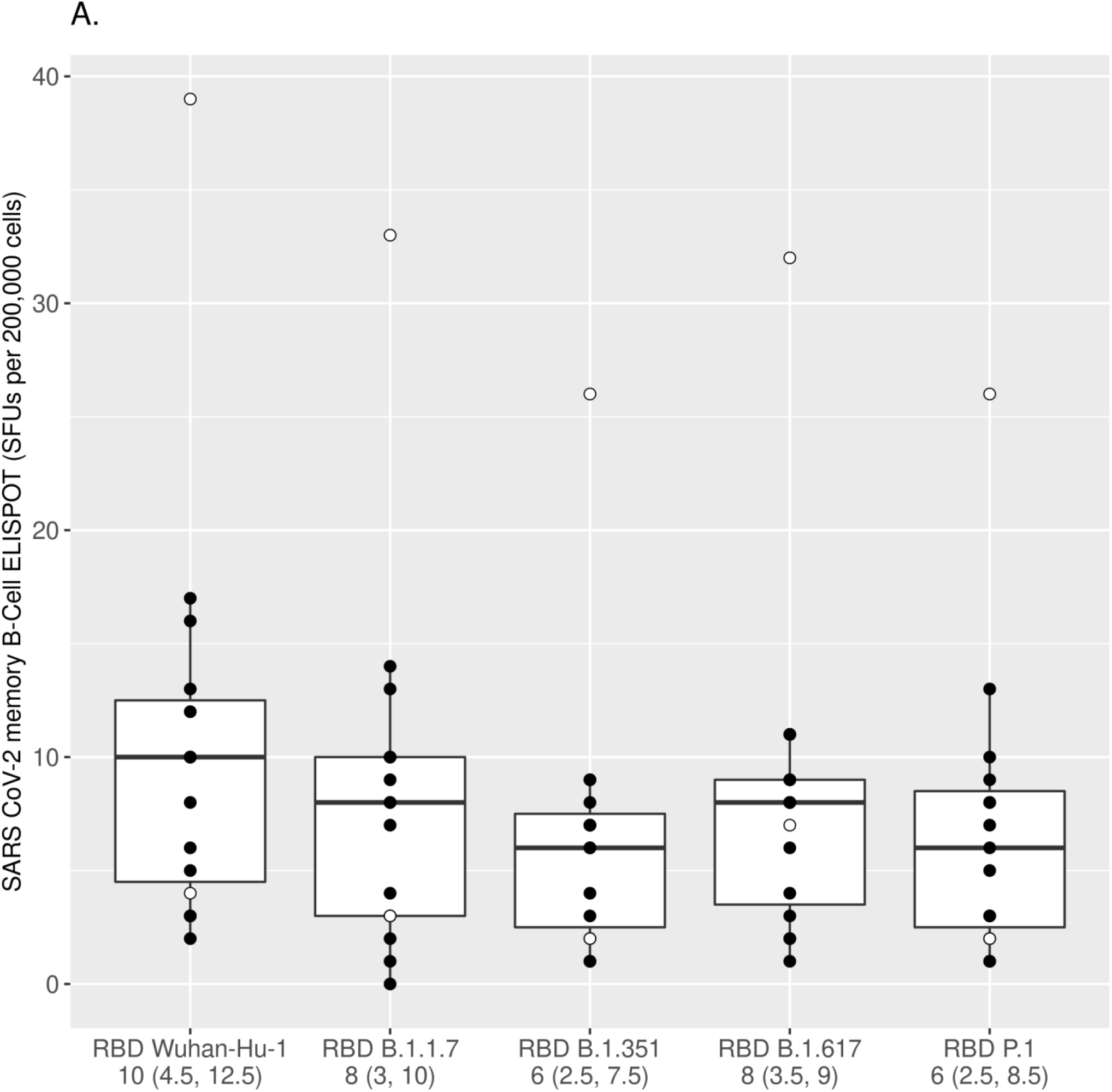
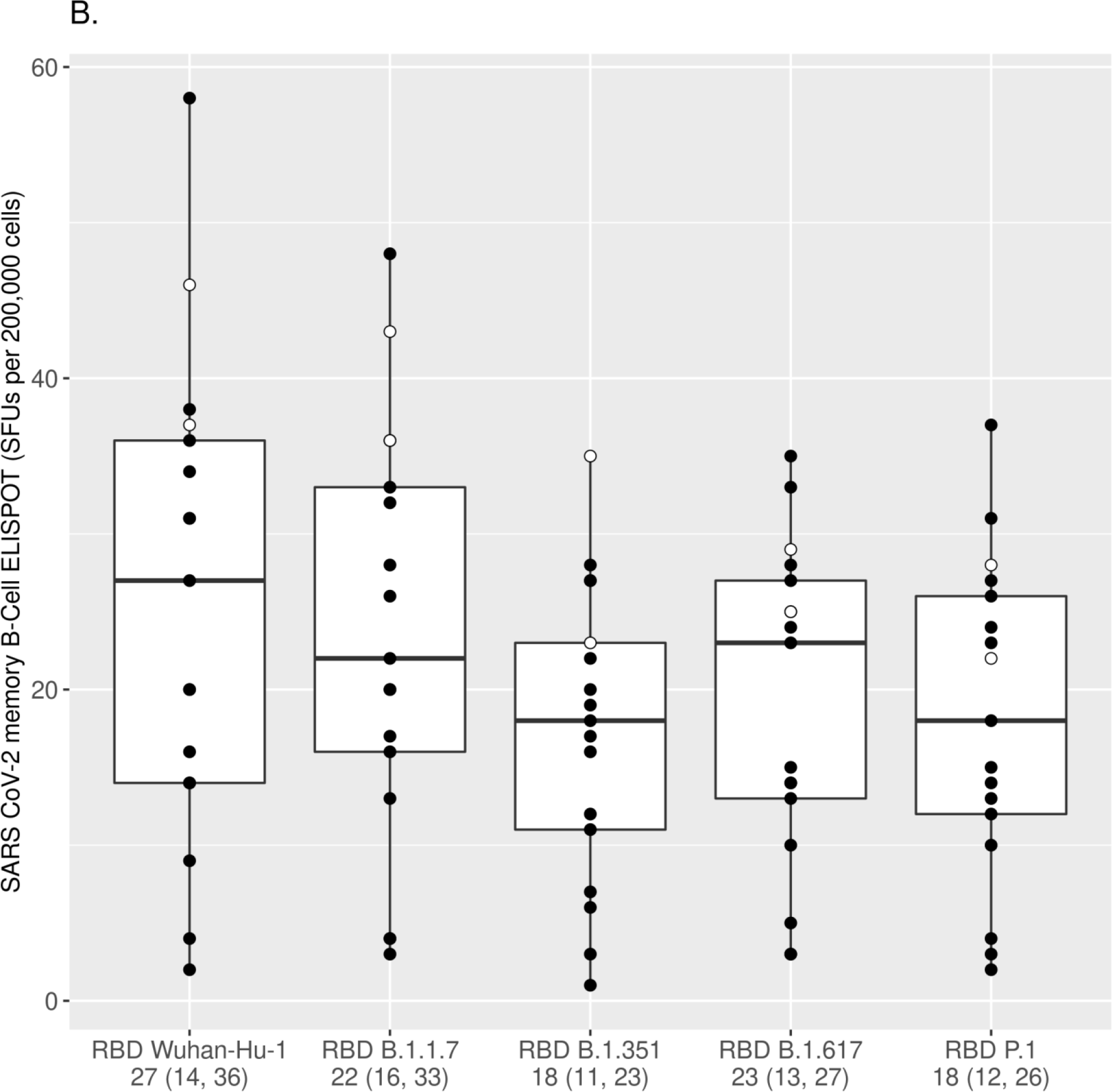
Reactivity of mRNA vaccine-induced MBCs to different VoC RBDs. The numbers of IgG+ MBCs reactive to the Wuhan-Hu-1 RBD and the RBDs of different VoC strains (Alpha/RBD B.1.1.7, Beta/RBD B.1.351, Gamma/RBD P.1 and Delta/RBD B.1.617) were measured using ELISPOT assay and assessed after the first mRNA vaccine dose (A) and after the second mRNA vaccine dose (B). Each box was plotted using the 25% to 75% interquartile range and the median was represented by the bold line in the box. The “ whiskers” extend up to 1.5 times the interquartile range above or below the 75th or 25th percentiles respectively. Black circles represent naïve (at baseline) subjects, while white circles represent COVID-19-recovered (at baseline) subjects. The numbers below the x-axis represent the median response for all subjects to the specified RBD antigen (25% and 75% interquartile range/IQR). The MBC response to each variant RBD was compared to the response directed to the Wuhan-Hu-1 RBD using the Wilcoxon signed-rank test [p<0.025 for all comparisons after the first dose (A); p<0.001 for all comparisons after the second dose (B) with the exception of Alpha/RBD B.1.1.7, which was non-significant after the second dose].

## Discussion

We and others have demonstrated the dynamics of SARS-CoV-2-specific antibody response after mRNA vaccination, and have demonstrated a reduced functional antibody response to emerging VoC.^2,5,6^ These findings corroborate the current understanding that COVID-19 vaccine-induced neutralizing antibody response may be less effective against newly arising VoC. Similar to recent findings, we found a robust S1/RBD-dominated MBC response after vaccination which benefits from a booster dose in COVID-19 naïve individuals and is correlated to a certain degree with antibody response. ^2^ A few studies assessed the SARS-CoV-2-specific MBC response after disease and determined the longevity of antigen-specific MBC response beyond six months after infection, as well as the similar MBC reactivity to one the Beta/RBD B.1.351.^1,7^ Our study builds on those findings by demonstrating (to the best of our knowledge), for the first time, differential specificity of mRNA vaccine-induced MBC response to multiple emerging VoC RBDs. While the MBC response to the VoC RBDs was consistently present after vaccination, we noted a statistically significant reduction in the number of isotype-switched/IgG+ memory B cells reactive to RBDs of Beta, Gamma and Delta SARS-CoV-2 variants compared to Wuhan-Hu-1 RBD. This, in concert with the lowered neutralizing antibody response to some of the variants, could translate to an increased susceptibility to emerging SARS-CoV-2 variant strains in the face of waning immunity. Without a correlate of protection the clinical significance of our findings remains uncertain at this time. These data further demonstrate the need for development and testing of broadly protective vaccines against SARS-CoV-2.^8^

## Data Availability

All data will be available regarding this study per request.

## Acknowledgments

We thank Nathaniel D. Warner for his help in statistical analyses. We thank Christina M. Mesa for editorial assistance. This study was supported by NIH grant (R01 AI48793: Poland PI) and Mayo Clinic funding provided to Dr. Kennedy for SARS-CoV-2 research.

## Conflict of Interest Statement

Dr. Poland is the chair of a Safety Evaluation Committee for novel investigational vaccine trials being conducted by Merck Research Laboratories. Dr. Poland offers consultative advice on vaccine development to Merck & Co. Inc., Avianax, Adjuvance, Valneva, Medicago, Sanofi Pasteur, GlaxoSmithKline, and Emergent Biosolutions. Drs. Poland and Ovsyannikova hold three patents related to measles and vaccinia peptide research. Dr. Kennedy holds a patent on vaccinia peptide research. Dr. Kennedy has received funding from Merck Research Laboratories to study waning immunity to measles and mumps after immunization with the MMR-II^®^ vaccine. Drs. Poland, Kennedy, and Ovsyannikova have received grant funding from ICW Ventures for preclinical studies on a peptide-based COVID-19 vaccine. All other authors declare no competing financial interests. This research has been reviewed by the Mayo Clinic Conflict of Interest Review Board and was conducted in compliance with Mayo Clinic Conflict of Interest policies.

## Notes

### Author Declarations

Study was approved by Mayo Clinic IRB #11-00017200

